# Pharmacologic and genetic evidence converge on mechanisms of psychotic illness

**DOI:** 10.1101/2024.04.30.24306623

**Authors:** Brian Fennessy, Liam Cotter, Nicole W. Simons, Lora E. Liharska, Girish N. Nadkarni, Douglas M. Ruderfer, Alexander W. Charney

## Abstract

Idiopathic and substance-induced forms of psychotic illness afflict millions of people worldwide, and it is largely unknown whether these two forms emerge through the same molecular mechanisms. Though genetic studies have implicated thousands of genes in idiopathic psychotic illnesses (e.g., schizophrenia), consensus is lacking regarding which of these genes are most likely to treat psychotic illness when modulated pharmacologically and, as a result, antipsychotic medications targeting these genes have yet to be developed. Previous studies suggest that one way to determine if a candidate target gene is likely to lead to an effective treatment for a given illness is if the gene is implicated by multiple lines of evidence (e.g., genetic, pharmacologic). Here, pharmacologic, genetic, and clinical data were leveraged to determine if the idiopathic and substance-induced forms of psychotic illness are related to one another through a common set of genes. A set of medications that cause psychotic illness as a side effect (“propsychotics”) were identified by analyzing 15 million medication side effects reports from over 100 countries. There was a significant overlap of target genes among propsychotics and antipsychotics and for many of the shared target genes propsychotics act through a mechanism that was qualitatively the opposite of the mechanism through which antipsychotics act (e.g., activation vs. inhibition). Propsychotic and antipsychotic target genes were significantly enriched for genes implicated in schizophrenia by rare loss-of-function genetic variation but not for genes implicated in schizophrenia by common genetic variation. Only one gene – *GRIN2A*, encoding the GluN2A subunit of the NMDA glutamate receptor – was implicated in psychotic illness by propsychotics, rare loss-of-function genetic variation, and common genetic variation. Mining genetic data from a diverse cohort of 30,000 adults treated in a New York City health system, a carrier of a rare loss-of-function variant in *GRIN2A* with severe psychotic illness was identified with a clinical course notable for psychotic symptoms and cognitive deficits that are not targeted by current antipsychotics. Altogether, this report shows how integrating pharmacologic, genetic, and clinical data from large cohorts can prioritize target genes for novel drug development and align the prioritized targets with specific clinical presentations.

## Introduction

The idiopathic and substance-induced forms of psychotic illness both afflict millions of people worldwide and are defined in the Diagnostic and Statistical Manual (DSM) by the presence of at least one of five types of psychotic symptoms: delusions, hallucinations, disorganized thinking, disorganized behavior, and negative symptoms^1^. Idiopathic forms include schizophrenia, schizoaffective disorder, and bipolar disorder. Substances that induce psychosis include amphetamines, phencyclidine, and psilocybin. The symptomatology and longitudinal course of psychotic illness can vary widely between affected individuals regardless of the cause^2–4^. Psychotic symptoms can be improved by “antipsychotics,” a class of medications that has been used in routine clinical practice to treat psychotic illnesses since the 1950s. Antipsychotics have helped millions of individuals and are featured in the list of essential medicines maintained by the World Health Organization to meet minimum needs of a basic health system^5^. Yet, antipsychotics are ineffective or intolerable in up to 75% of affected individuals, do not treat all the symptoms of psychotic illness, and do not modify disease progression^6–9^. The primary molecular mechanism through which most antipsychotics are believed to exert clinical effects – dopamine receptor antagonism – has remained unchanged since the first antipsychotic chlorpromazine was introduced. Therefore, a long-standing and urgent unmet need is the development of a new generation of antipsychotics.

To develop a new generation of antipsychotics, target genes (i.e., the gene products through which a medication exerts clinical effects) must be prioritized for further investigation. A proven strategy for prioritizing target genes is to leverage knowledge of the pharmacology of psychotic illness (i.e., knowledge of the mechanisms through which substances modulate symptoms of psychotic illness). Dopamine receptors, serotonin receptors, and muscarinic receptors – which, collectively, comprise the target genes of most antipsychotics used in clinical practice – were all prioritized as target genes based on observations that psychotic symptoms can be induced and/or treated through pharmacological modulation of these receptors^10–12^. Using knowledge of pharmacology to prioritize target genes for drug development is now a scalable research strategy, as large databases linking medications to clinical effects and target genes have been created that can be mined using computational techniques. Of the many target genes that can be prioritized using these techniques, only a small number are likely to result in the development of effective treatments. Previous studies suggest that one way to determine if a target gene is likely to lead to an effective treatment is to consider whether genetic variation linked to that gene contributes to increased risk of that illness^13^.

The most common idiopathic forms of psychotic illness (e.g., schizophrenia) are up to 80% heritable^14^, and as such these illnesses have been a major focus of human genetics research. Studies of large populations have identified many genetic variants that contribute to schizophrenia and other idiopathic forms of psychotic illness (“psychosis risk variants”), and these variants are linked to thousands of genes (“psychosis risk genes”). The class of psychosis risk variants that make the greatest contribution to the heritability of psychotic illnesses are common single-nucleotide polymorphisms (SNPs), which account for up to ∼25% of the heritability of schizophrenia^15^. Risk variants of this class are identified through genome-wide association studies (GWAS), and the most recent schizophrenia GWAS – the third GWAS reported by the Psychiatric Genomics Consortium Schizophrenia Working Group (“the PGC3SCZ GWAS”) – identified risk SNPs in 287 genomic regions including thousands of genes^15^. Rare loss-of-function (LoF) variants comprise another class of psychosis risk variants, and these are variants that are rare in the population and change the coding sequence of a gene in a manner predicted to result in a dysfunctional gene product^16,17^. Recently, the Schizophrenia Exome Meta-analysis (SCHEMA) study reported 10 genes harboring an excess of rare LoF variants in cases compared to controls^17^ and a subsequent study identified two additional genes^16^. Rare LoF variants account for a small amount of the heritability of psychotic illnesses but can contribute substantially to risk in individual cases. While thousands of genes have been implicated in psychotic illness through pharmacology and genetics research only a few antipsychotics have been developed that target these genes^18^, in part because there is a lack of consensus regarding which of the many genes implicated should be pursued as a target of novel treatments.

Here, pharmacologic, genetic, and clinical data are leveraged to prioritize target genes for novel antipsychotic development. A large database of medication side effect reports is used to identify medications that cause psychotic illness as a side effect (“propsychotics”). Using databases that link medications to target genes, propsychotic target genes are identified and compared to antipsychotic target genes, revealing a shared set of targets that propsychotics act on through a mechanism (e.g., activation) that is qualitatively the opposite of the mechanism exerted by antipsychotics on the targets (e.g., inhibition). Significant overlap was observed between propsychotic target genes and genes implicated in psychotic illnesses by rare LoF variants but not between propsychotic target genes and genes implicated in psychotic illnesses by common SNPs. By aggregating pharmacologic and genetic data, activation of *GRIN2A* is prioritized as a mechanism to pursue in the development of novel antipsychotics. To begin to determine the clinical presentation of psychotic illness that may be most likely to respond to this pharmacologic mechanism, a case report of schizophrenia linked to a rare LoF variant in *GRIN2A* is presented that is most notable for the prominence of disorganized thought, disorganized behavior, deficits in cognitive function, and co-morbid epilepsy. Altogether, this report provides an approach to prioritize individual target genes to pursue in developing novel treatments for a highly polygenic and symptomatically heterogeneous illness.

## Results

### Defining propsychotics

For each database used in the current report to link medications to clinical effects (i.e., side effects, indications) or target genes, the medication and clinical effect terms used in the database were standardized to RxNorm and Medical Dictionary for Regulatory Activities [MedDRA] terms, respectively (***Supplementary Information***). Psychosis side effects were defined as a manually curated set of MedDRA terms relevant to psychosis. VigiBase^19^, a medication side effect reporting database with over 15 million reports maintained by the World Health Organization (WHO), was used to identify medications that induce psychotic symptoms as a side effect (i.e., propsychotics) (***Figure 1***). Each VigiBase report represents an instance where a medical professional suspects a medication has caused a side effect. The statistical significance of each reported medication side effect in VigiBase was assessed using disproportionality analysis, which tests if the medication and side effect co-occur in the reports of the database more than expected by chance^20,21^. After excluding antipsychotics from consideration (***Supplementary Table 1***), 276 medications were defined as propsychotics by being linked to >= 1 psychosis side effect term (66 psychosis side effect terms were linked to >= 1 propsychotic; ***Supplementary Tables 2-3***). The psychosis side effect terms linked to the greatest number of propsychotics were hallucinations (linked to 111 propsychotics), psychotic disorder (linked to 61 propsychotics), visual hallucinations (linked to 54 propsychotics), depersonalization/derealization disorder (linked to 48 propsychotics), and paranoia (linked to 35 propsychotics) (***Figure 2; Supplementary Table 4***). Using ATC level 3 categories to group medications into pharmacological subgroups, propsychotics were found to span 85 different pharmacological subgroups. The five subgroups that included the most propsychotics were hypnotics and sedatives (16 propsychotics), antihistamines (15 propsychotics), antidepressants (15 propsychotics), antiepileptics (13 propsychotics), and dopaminergic agents (12 propsychotics) (***Figure 3; Supplementary Table 5***). The Side Effect Resource (SIDER), a database that links medications to side effects reported on Food and Drug Administration (FDA) labels^22^, was used to validate the set of 276 propsychotics defined using VigiBase. Of the 1,334 unique medications in SIDER, 412 were linked to at least one psychosis side effect term and were not an antipsychotic. The number of medications overlapping this set and the set of propsychotics identified using VigiBase (N = 148 in both sets) represented a 2.57-fold increase of the number expected by chance (hypergeometric p-value = 1.15 x 10^-46^).

**Figure 1.**
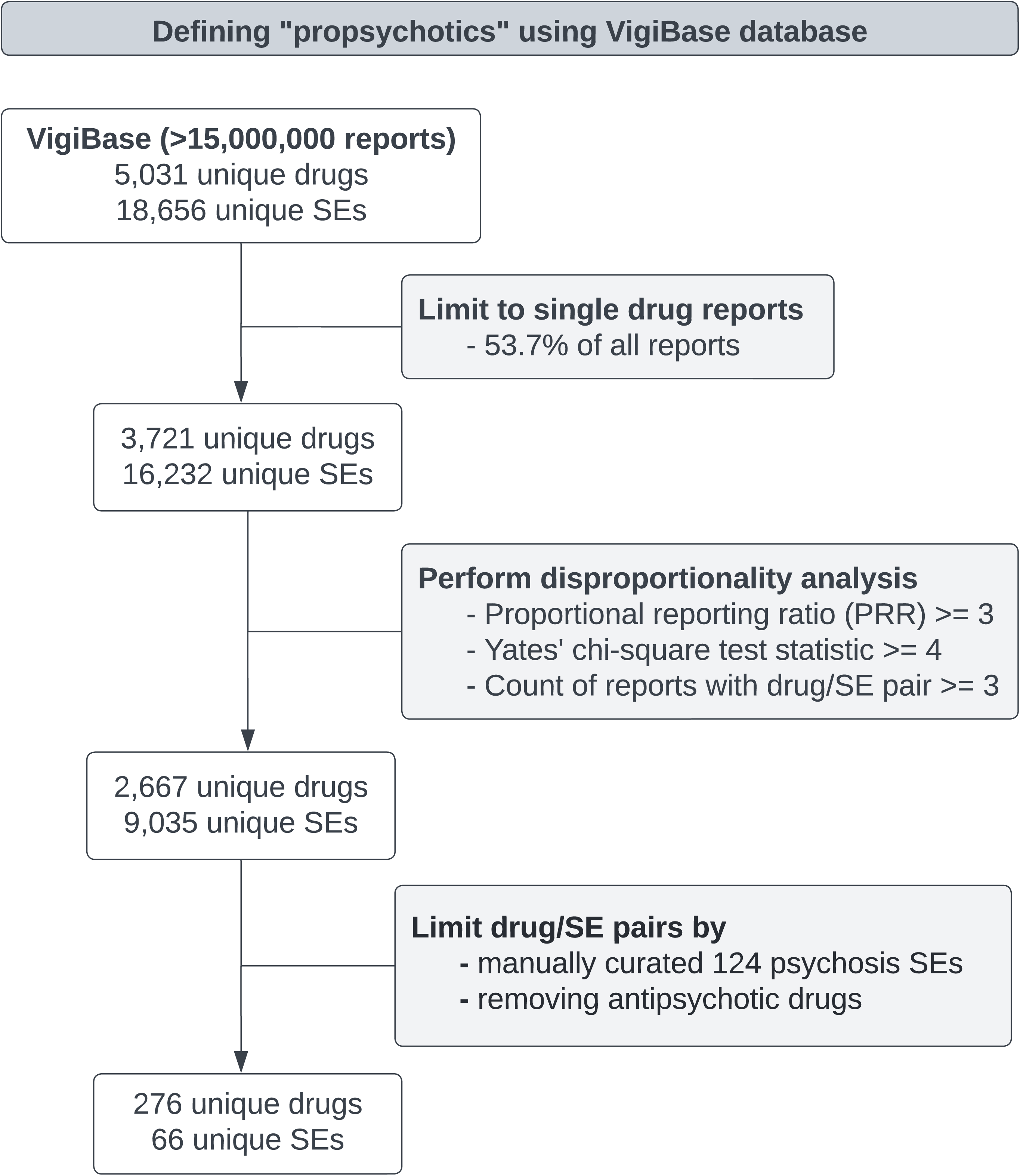
Summary of the workflow used to define propsychotics in VigiBase. SE: side effect.

**Figure 2.**
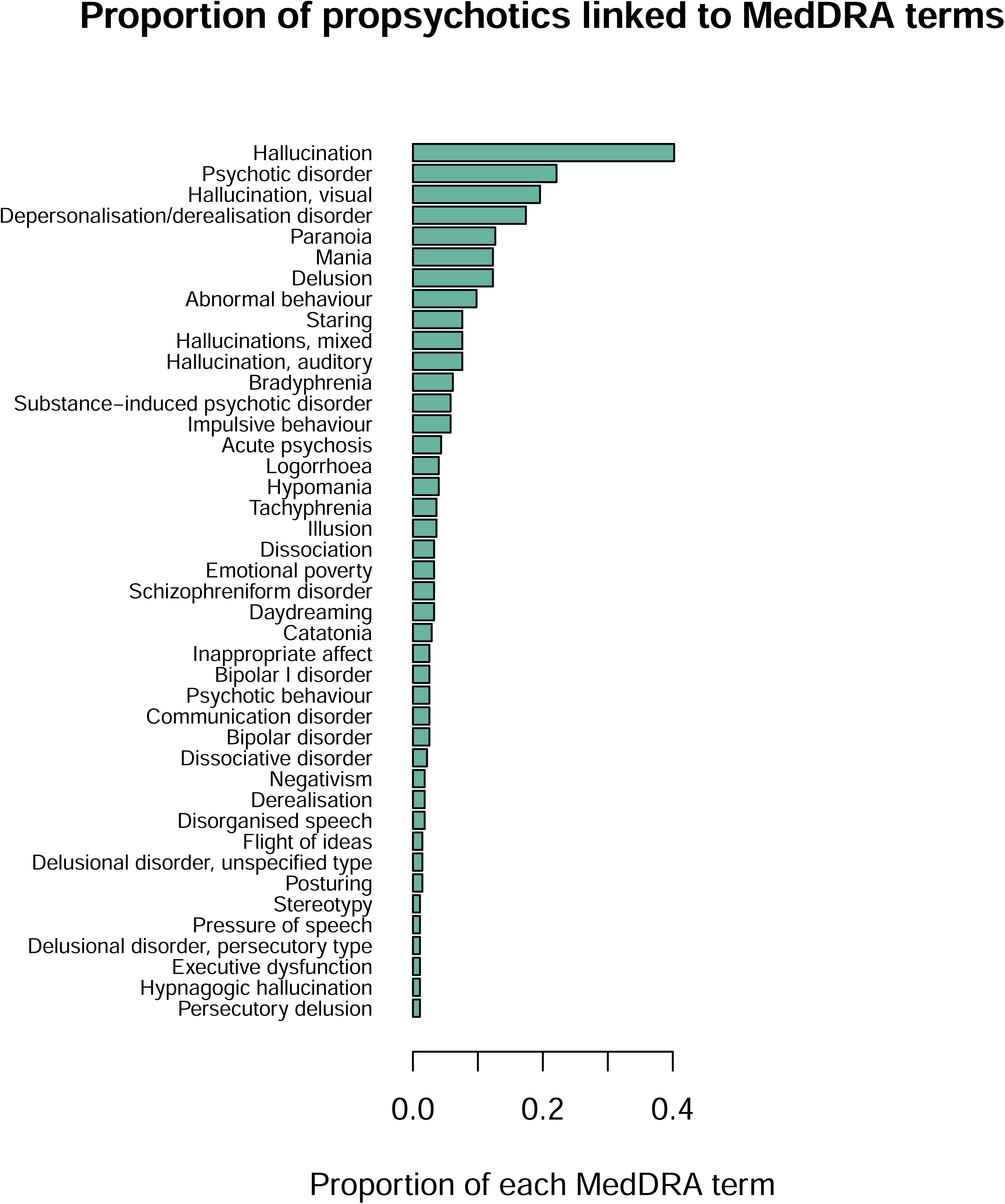
Proportion of propsychotics linked to MedDRA terms. The vertical axis shows MedDRA psychosis side effect terms. The horizontal axis shows the proportion of propsychotics that were linked to the MedDRA psychosis side effect term on the vertical axis. Only MedDRA psychosis side effect terms linked to greater than 1% of propsychotics are shown.

**Figure 3.**
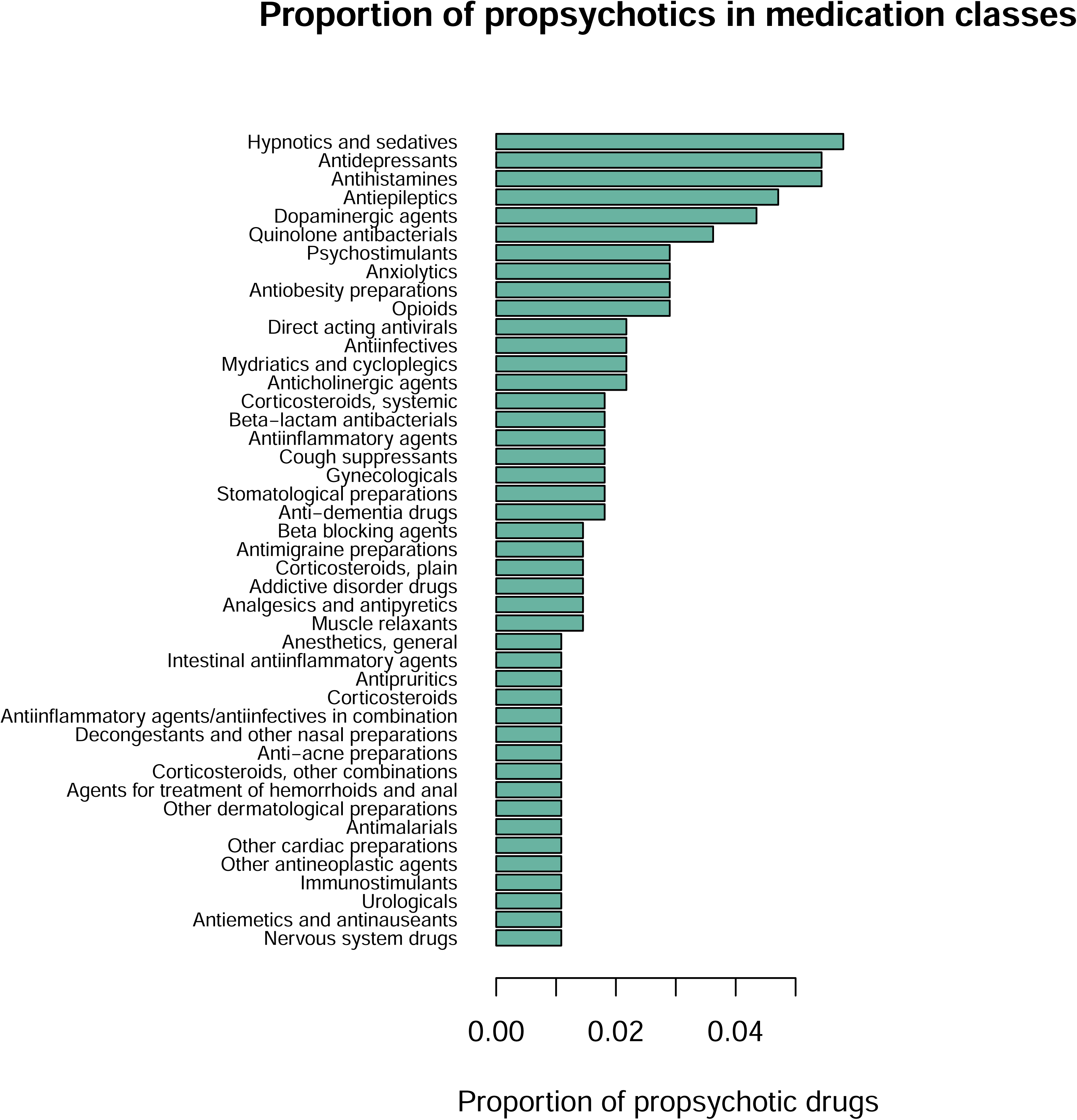
**Proportion of propsychotics in medication classes. The vertical axis shows ATC Level 5 medication class. The horizontal axis shows the proportion of propsychotics that were linked to the ATC Level 5 medication class on the vertical axis. Only ATC classes linked to greater than 1% of propsychotics are shown.**

### Comparing mechanisms of propsychotics and antipsychotics

Two databases were used to link medications to target genes: (1) DrugBank, which links medications to target genes through curation of scientific literature^23^; (2) SeaChange, which predicts the target genes of a medication based on the chemical structure of the medication^24^. Of the 276 propsychotics identified in VigiBase, 240 were linked to at least one target gene through either DrugBank or SeaChange (1,134 target genes were linked to at least one propsychotic). A permutation approach was used to identify genes significantly overrepresented as targets of propsychotics. For each putative target gene, the number of propsychotics targeting the gene was compared to the number of medications targeting the gene in a randomly selected set of 240 medications. This analysis yielded 170 propsychotic target genes with an empirical p-value below 0.05 after 100,000 permutations (***Supplementary Table 6***). Using a similar procedure to define propsychotic target genes, 129 antipsychotic target genes were identified with an empirical p-value below 0.05 after 100,000 permutations (***Supplementary Table 6***). A significant overlap was observed between the 170 propsychotic target genes and the 129 antipsychotic target genes (N = 67 shared target genes; Fisher’s exact test odds ratio [OR] = 33.3, p-value = 1.57 x 10^-57^). This overlap remained significant when considering (1) only experimentally validated target genes defined in DrugBank (OR = 110.0, p-value = 2.0 x 10^-34^) and (2) only propsychotics not classified as nervous system medications (OR = 22.9, p-value = 1.61 x 10^-38^).

DrugBank contains data on the mechanisms of action of medications on target genes (i.e., whether a medication activates or inhibits the activity of a target gene). Each mechanism of action term in DrugBank (e.g., “agonist”) was classified as either an activating action or an inhibiting action. Mechanism of action information was available for 107 of the 170 propsychotic target genes and for 45 of the 129 antipsychotic target genes. For both the propsychotic and antipsychotic medication classes, a permutation procedure was used to determine if for a given target gene the medications in the class exerted either an activating or inhibiting action more than expected by chance (i.e., more than was observed for random sets of medications chosen over 100,000 permutations; ***Supplementary Table 7***). Propsychotics were found to exert an activating action on 52 target genes and an inhibiting action on 63 target genes (for 25 target genes, propsychotics were found to exert both an activating and inhibiting action). Antipsychotics were found to exert an activating action on 6 target genes and an inhibiting action on 45 target genes. As expected, the target gene inhibited by the largest number of antipsychotics is *DRD2* (inhibited by over 75% of antipsychotics, ***Figure 4A***). For 24 of the genes targeted by both propsychotics and antipsychotics (“shared target genes”), propsychotics were found to act on the target through a mechanism that is qualitatively the opposite of the mechanism through which antipsychotics were found to act on the target (***Figure 4A***). Target genes were grouped by neurotransmitter receptor class (***Figure 4A***), revealing for dopaminergic, serotonergic, muscarinic, and adrenergic receptor classes propsychotics exert activating actions while antipsychotics exert inhibiting actions (***Supplementary Table 8***). For several receptor classes, a significant mechanism of action was found for propsychotics but not for antipsychotics, and these include gamma-aminobutyric acid (GABA) and glutamate receptors, upon which propsychotics exert activating and inhibiting actions, respectively (***Figure 4; Supplementary Table 8***). Propsychotics and antipsychotics exert the same action on 27 target genes, the majority of which (N = 20) are also amongst the 24 target genes where propsychotics and antipsychotics exerted opposing actions.

**Figure 4.**
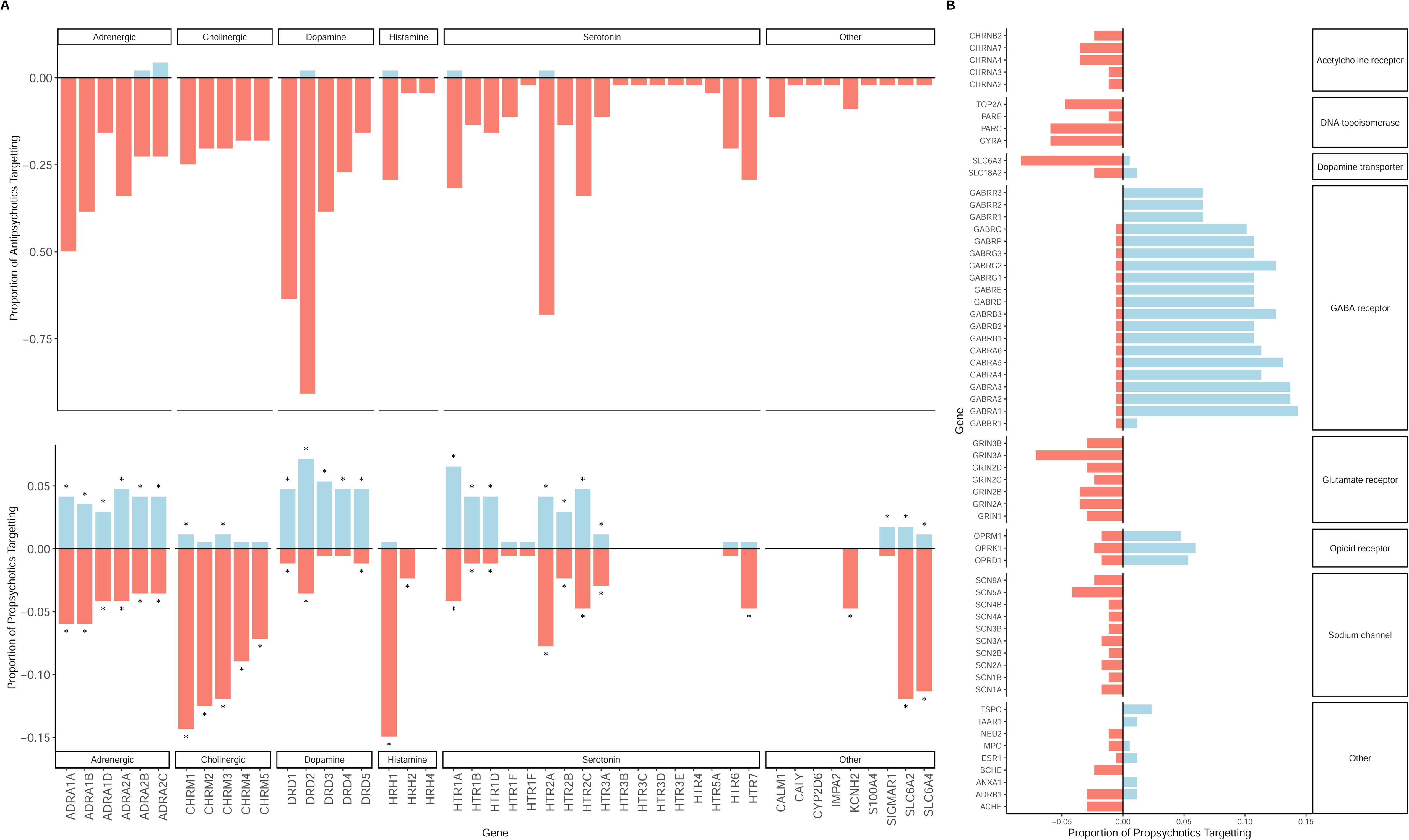
(A) For every significant antipsychotic target gene mechanism of action there is an entry on the x-axis. The top panel shows on the y-axis the proportion of antipsychotics that exerted an activating mechanism of action on the target gene (blue) and the proportion of antipsychotics that exerted an inhibiting mechanism of action on the target gene (orange; represented as negative values), and only significant values are shown. The bottom panel shows on the y-axis the proportion of propsychotics that exerted an activating mechanism of action on the target gene (blue) and the proportion of antipsychotics that exerted an inhibiting mechanism of action on the target gene (orange; represented as negative values), and significant values are indicated by an asterisk. Target genes are grouped by neurotransmitter receptor class as indicated by the facet labels. (B) For every significant propsychotic mechanism of action that did not have a significant mechanism of action for antipsychotics there is an entry on the y-axis. On the x-axis is shown the proportion of propsychotics that exerted an activating mechanism of action on the target gene (blue) and the proportion of antipsychotics that exerted an inhibiting mechanism of action on the target gene (orange; represented as negative values), and only significant values are shown.

### Enrichment of propsychotic and antipsychotic target genes for psychosis risk genes

Analyses were performed testing whether propsychotic and antipsychotic target genes are enriched for genes implicated in psychotic illness through large-scale genetic studies. The strength of association between psychosis risk and a gene was defined in two ways: (1) “rare LoF variant psychosis risk” was defined as the p-value for association between the burden of rare LoF variants in the gene and schizophrenia in the SCHEMA study^17^; (2) “common SNP psychosis risk” was defined as the p-value for association between the gene and schizophrenia (defined using MAGMA) in the PGC3SCZ GWAS^15^. Only genes that were a target of at least one medication in the databases used in this report were included in these analyses. For antipsychotics and propsychotics, two two-sample Wilcoxon tests were run that each compared target genes to non-target genes – one test to assess if target genes had greater rare LoF psychosis risk and another test to assess if target genes had greater common SNP psychosis risk. Confirming findings from prior work^25^, antipsychotic target genes had increased rare LoF psychosis risk compared to other genes (p-value = 0.006) but no difference in common SNP psychosis risk compared to other genes (p-value = 0.08). Similarly, propsychotic target genes had increased rare LoF psychosis risk compared to other genes (p-value = 0.011) and no difference in common SNP psychosis risk compared to other genes (p-value = 0.23). High-confidence psychosis risk genes were defined as genes with statistically significant associations with schizophrenia in either the SCHEMA study (N = 10 as defined by the SCHEMA authors; “rare LoF psychosis risk genes”) or the PGC3SCZ GWAS (N = 636 with significant MAGMA p-values after multiple test correction; “common SNP psychosis risk genes”). Among the 170 propsychotic target genes, 8 were a high-confidence psychosis risk gene (*GRIN2A*, *DRD2*, *CYP2D6*, *CHRNB4*, *CHRM4*, *GABBR1*, *GRM3*, *CHRNA3*). All 8 of these were a common SNP psychosis risk gene, but only a single gene – *GRIN2A* – was implicated in psychotic illness as a propsychotic target gene (***Figure 4B***), a rare LoF psychosis risk gene, and a common SNP psychosis risk gene.

### Clinical characteristics of psychotic illness in a carrier of a rare LoF variant in *GRIN2A*

*GRIN2A* was associated with schizophrenia through rare LoF variants in the SCHEMA study, but the clinical presentations of the schizophrenia cases harboring these variants in that study were not described^26^. The results in the previous section suggest activation of *GRIN2A* is a promising mechanism to pursue in developing novel antipsychotics, and since schizophrenia is a heterogeneous clinical condition, it is possible that activation of *GRIN2A* may be most promising for treating individuals with a specific clinical presentation. As a first step towards characterizing the schizophrenia clinical presentation that may be most likely to improve with activation of *GRIN2A*, whole-exome sequencing data from approximately 30,000 individuals in a large United States health system were mined to identify and clinically characterize carriers of rare LoF variants in *GRIN2A* (i.e., using the same definitions of rare and of LoF used in the study that implicated *GRIN2A* rare LoF variants in schizophrenia). One carrier was identified who had a previous diagnosis of schizophrenia, and the rare LoF variant in this carrier (“the founder”) is located at position 9,798,422 on chromosome 16 within the exon ENSE00001304023 (genome build GRCh38/hg38). The founder is a heterozygote at this position, with one copy of the reference allele G and one copy of the alternate allele C. The alternate allele C has a frequency of 0 in the Genome Aggregation Database (gnomAD; v4.0.0). In the coding sequence of *GRIN2A*, the presence of this variant results in a premature stop codon in four of the known RNA transcript isoforms of *GRIN2A* (ENST00000330684, ENST00000396573, ENST00000535259, and ENST00000562109). A clinical case history was assembled through chart review and interviews with the founder and the founder’s family members. Four notable observations emerged from this case history. First, the onset of neurological and mental illness in the founder was in childhood, when the founder was diagnosed with a seizure disorder and a non-specific intellectual disability referred to by treating physicians in the medical chart as “mild mental retardation.” Second, in adolescence, the intellectual disability persisted, as evidenced by the founder’s placement first in a high school and later in a job training program designated for individuals with such disabilities. Third, when the founder reached the fourth decade of life, psychosis came to dominate the clinical picture with the first of seven inpatient hospitalizations for severe psychosis. Of the five domains of psychotic illness defined in the DSM, the acute psychotic presentations documented for the founder were most notable for disorganized thinking (e.g., loosening of associations, incomprehensible speech) and disorganized behavior (e.g., unpredictable agitation, going missing from home, inappropriate laughter). The founder has never experienced hallucinations or delusions, and the negative symptoms of psychosis (e.g., avolition, anhedonia) – while not absent altogether – seem to have been moderate. Fourth, the founder has multiple siblings who also reportedly have medical histories notable for psychotic illnesses, seizure disorders, and/or non-specific intellectual disabilities (genotypes and detailed case histories are not available from these individuals at the time of writing).

## Discussion

This report integrates pharmacologic, genetic, and clinical data from a total of over 15 million individuals to make four key observations. First, a set of medications linked to psychotic illness as a side effect were defined (propsychotics). Second, the target genes of propsychotics were found to overlap with the target genes of antipsychotics, and for many of these shared target genes the action exerted by propsychotics on the gene (e.g., activation) was found to be the opposite of the action exerted by antipsychotics on the gene (e.g., inhibition). Third, both propsychotic and antipsychotic target genes were found to be enriched for genes implicated in schizophrenia through rare loss-of-function genetic variation but not through common genetic variation. Fourth, activation of *GRIN2A* was prioritized as a mechanism to develop new antipsychotics around, and a clinical case report of schizophrenia in a carrier of a rare LoF variant in *GRIN2A* provided clues regarding the schizophrenia clinical presentation to target with this pharmacological strategy.

Propsychotics were defined by mining a large-scale medication side effect reporting database. These medications spanned many classes, including some that were intuitively expected (e.g., classes of medications that were designed to act in the nervous system [e.g., dopaminergic agents]) and some that were not (e.g., quinolone antibiotics). The set of propsychotics defined using this approach was validated using independent data, and some of the less intuitive medication classes represented in propsychotics had prior literature support for association to psychotic illness^27^. Since VigiBase is limited to reports about medications that are used in clinical practice, propsychotics defined in this report do not include substances that are used to induce psychotic experiences recreationally (e.g., PCP, MDMA, psilocybin). In recent years, several of these substances have become treatments of affective illnesses^28–30^ but these agents were not represented by reports in the version of VigiBase analyzed for this report and therefore are not driving the primary findings of the study.

The approach used to link propsychotics to target genes highlights one way to systematically identify genes capable of inducing a complex illness in humans when acted upon pharmacologically. The 170 propsychotic target genes are highly overlapping with the 129 antipsychotic target genes found using this approach, and for many of the genes targeted by both propsychotics and antipsychotics (“shared target genes”), propsychotics act on the target through a mechanism that is qualitatively the opposite of the mechanism through which antipsychotics act on the target. While for a few of the shared target genes this observation is not novel^12^, for most shared target genes there are no previous reports of the observation. Receptors from most of the major neurotransmitter systems are represented in both propsychotic and antipsychotic target genes, illustrating the complexity of psychosis pharmacology and supporting the notion that there are multiple pharmacological mechanisms through which psychotic illness can be both induced and treated. Interestingly, both antipsychotics and propsychotics were found to exhibit inhibiting actions on *CHRM4* (***Figure 4A***), which encodes the muscarinic acetylcholine receptor M_4_ and is the primary target gene of the newly-approved antipsychotic xanomeline that exerts an activating action on *CHRM4*^31^ (since xanomeline was just recently approved for clinical use, it is not in the set of antipsychotics defined by the ATC used in this report). As expected, over 75% of antipsychotics exert inhibiting actions on *DRD2* and over 50% of antipsychotics exert inhibiting actions on *HTR2A*; in contrast, the percentage of propsychotics that act on a target gene through the same mechanism is less than 15% for all target genes. This difference highlights that the method used to define propsychotics for this report did not simply identify a set of medications that act through a mechanism that is the inverse of the primary mechanism of antipsychotics. Indeed, receptors for several neurotransmitter classes represented in propsychotic target genes are not represented in antipsychotic target genes, including receptors for GABA (i.e., the primary inhibitory neurotransmitter in the central nervous system) and glutamate (i.e., the primary excitatory neurotransmitter in the central nervous system). The observation that propsychotics primarily activate GABA receptors and primarily inhibit glutamate receptors suggests that (1) a state of psychotic illness can be induced pharmacologically in an individual either by increasing inhibitory tone or by decreasing excitatory tone and (2) maintaining a balance between excitatory and inhibitory tone may be key for maintaining an individual in a non-psychotic state.

The complex picture of psychosis pharmacology that emerged from linking propsychotics to their target genes (i.e., the large number of target genes that included receptors for many neurotransmitter classes) appeared consistent with the complex picture of the genetic architecture of psychotic illness that has been emerging from population genetics over the past two decades. When this apparent consistency was more deeply investigated, the target genes of propsychotics were found to be enriched for genes implicated in psychotic illness through rare loss-of-function genetic variation but not for genes implicated in psychotic illness through common genetic variation. This finding is consistent with an earlier study of antipsychotics only^25^ and validates the notion that propsychotic target genes contribute to psychotic symptoms. Common genetic variation accounts for a substantial amount of the heritability of psychotic illnesses at the population level, while rare loss-of-function genetic variation accounts for a small amount of the heritability at the population level but can contribute substantially to risk at the individual level. The observation that propsychotic target genes overlap with the genes linked to psychosis through rare loss-of-function genetic variation may reflect that there is a relatively small subset of genes in the human genome capable of exerting large effects on psychosis risk when dysfunctional. These genes, which can be identified by as the genes implicated in psychotic illness through both pharmacologic and genetic evidence, may be the most promising candidate target genes for novel antipsychotics^13^.

*GRIN2A* is a gene located in chromosome 16 that encodes for a subunit of the N-methyl-D-aspartate (NMDA)-type glutamate receptor (NMDAR) and was the only gene in the current report linked to psychotic illness as a propsychotic target gene, a rare LoF psychosis risk gene, and a common SNP psychosis risk gene. NMDARs are ionotropic glutamate receptors involved in many aspects of excitatory neurotransmission^32^. Each NMDAR is composed of four protein subunits that, together, span the plasma membrane and form an ion channel pore, and the biological properties of NMDARs are determined by the combination of subunits used. The proteins GluN1, GluN2, and GluN3 are the possible NMDAR subunits, and every NMDAR is composed of two GluN1 subunits with (1) two GluN2 subunits, (2) two GluN3 subunits, or (3) one GluN2 subunit and one GluN3 subunit. The GluN2A subunit encoded by GRIN2A is one of four distinct types of GluN2 subunits (the other three are GluN2B, GluN2C, and GluN2D), each encoded by a different gene. The biological processes that rely on GluN2A-containing NMDARs are still a matter of debate but may include long-term potentiation, a type of synaptic plasticity where synaptic activity induces a chronic increase in signal transmission between two neurons^33^. *GRIN2A* is one of only two genes in the genome that have been linked to schizophrenia through both rare loss-of-function genetic variation and common genetic variation, and the other gene, *SP4*, encodes a transcription factor that regulates *GRIN2A* expression^1,2,4^. Decreased NMDAR activity is suggested as a mechanism in the pathogenesis of psychotic illness not only by the pharmacologic and genetic evidence presented in this report, but also by (1) clinical immunology, where autoantibodies against NMDAR subunits (including the GluN2A subunit) downregulate NMDAR activity and cause a psychotic illness that clinically can be indistinguishable from schizophrenia^6^ and (2) rodent models of NMDA receptor hypofunction, where restoring NMDA receptor activity rescues psychosis-like phenotypes^7^. Taken together, these diverse lines of evidence support the hypothesis that pharmacologically increasing *GRIN2A* activity could treat psychotic symptoms. This hypothesis has not been adequately tested in human clinical trials.

In addition to identifying target genes, identifying clinical presentations to target is key for novel antipsychotic development^34^. The clinical case summary in this report of a carrier of a rare loss-of-function variant affected with schizophrenia begins to seek out the presentations of psychotic illness that may be most likely to respond to pharmacologic activation of *GRIN2A*. This is not the first case summary to characterize carriers of putatively disease-causing genetic variants in *GRIN2A*^35–37^. What separates the case described here, however, is the approach used to identify the carrier: the variant annotation strategy was modeled closely after the variant annotation strategy used in the study that linked rare loss-of-function variation in *GRIN2A* to schizophrenia at the population level (i.e. the SCHEMA study)^17^. The most prominent symptoms of the illness in this individual (e.g., disorganized thought and behavior, cognitive deficits) are amongst the symptoms of schizophrenia that are the most debilitating and the least responsive to current antipsychotics. In addition to schizophrenia, large-scale genetic studies have also implicated rare loss-of-function variants in *GRIN2A* to seizure and neurodevelopmental disorders, and it has been proposed that the illness that results from a rare loss-of-function variants in *GRIN2A* (i.e., schizophrenia vs. epilepsy vs. intellectual disability) depends on the specific loss-of-function variant^37^. However, the clinical course reported here – which included schizophrenia, epilepsy, and intellectual disability all in the same individual – suggests the same loss-of-function variant may predispose to all of these illnesses. As novel antipsychotics that activate *GRIN2A* are developed, clinical trials may be more likely to success if the participants included are enriched for those with similar clinical presentations to the case presented in this report.

There are several limitations to this work. First, the propsychotics and propsychotic target genes were identified based on the set of MedDRA terms used to define psychosis. This set of terms was manually constructed by a single study psychiatrist, and other experts when presented with the same task may have arrived at a different set of terms. Second, for most propsychotics defined by this report psychosis side effects are rare, so use of these medications to model psychosis in future research may be challenging. Third, the data linking medications to target genes and mechanisms of action is incomplete, simplified for interpretation, and/or lacking with respect to key variables such as binding affinity. Fourth, knowledge of the genetic architecture of psychotic illnesses is evolving, and it is possible that some of the findings in this report may change as the list of specific genes implicated in psychotic illness expands in the future. Fifth, the clinical presentation of only a single case of schizophrenia in a carrier of a rare loss-of-function variant in *GRIN2A* is examined. As such, the features of the clinical presentation may not be typical of all *GRIN2A*-linked cases of schizophrenia and larger cohort studies are needed to validate the observations from this single case and more rigorously delineate the clinical presentation associated with dysfunction in this gene.

Historically, whether idiopathic or substance-induced forms of a disease emerge through the same molecular mechanisms has been inadequately studied. This report approaches this question with a focus on psychotic illness and finds that a select set of genes are implicated through both genetics and pharmacology. These genes should be prioritized as target genes for developing a new generation of antipsychotics, with a particular emphasis on activation of *GRIN2A* given the multiple lines of evidence that together show downregulation of this gene induces psychotic illness in humans. Clinical characterization of psychotic illness in individuals with rare loss-of-function variants in genes such as *GRIN2A* will be crucial to link symptom presentation to potential therapeutic benefit. This framework provides a pathway to systematically prioritize genetically supported target genes for developing novel treatments for diseases that are polygenic and symptomatically heterogeneous.

## Methods

### Standardizing medication names and clinical concepts

The databases utilized for this study to link medications to clinical concepts (i.e., side effects, indications; VigiBase, SIDER) and target genes (i.e., DrugBank, SeaChange) used a variety of lexicons to identify medications and clinical concepts. To analyze the databases together, names used for medications had to be mapped to a single lexicon and names used for clinical concepts had to be mapped to a single lexicon. Detailed descriptions of the procedures used to standardize medication and clinical concept names are provided in the ***Supplementary Information*** and described in brief here. Medication names were standardized to RxNorm (version of September 4th, 2018) Concept Unique Identifiers (RXCUIs) with a Source Abbreviation (SAB) value of “RXNORM” and a Term Type (TTY) of “IN.” Clinical concept names were standardized to the Medical Dictionary for Regulatory Activities (MedDRA; version 20.0) Preferred Terms (PT). For analyses that required medications to be categorized into medication classes, the primary database used to categorize medications was the World Health Organization Anatomical Therapeutic Chemical Classification (ATC)^38^. ATC organizes medications into a hierarchy of five levels, and ATC Level 3 (e.g., “Antipsychotics”) was for this study used to categorize medications into classes based on physiological mechanisms and therapeutic properties. For analyses that required all medications targeting the nervous system to be defined, the ATC Level 1 value “Nervous System” was used.

### Defining psychosis side effect terms

Multiple steps in the study (e.g., defining propsychotics) required the broad clinical concept of psychotic illness to be defined in the MedDRA lexicon terms. Since there is no single MedDRA PT that fully captures the complexity of psychotic illness, a set of MedDRA PTs was manually defined for the current study to define psychotic illness in MedDRA terms. A study psychiatrist (AWC) manually reviewed each of the approximately 500 PTs under the MedDRA System Organ Class “Psychiatric Disorder” and based on clinical experience identified the subset of these PTs that described either a psychotic symptom (e.g., “Hallucinations”) or a psychotic syndrome (e.g., “psychotic disorder”). This manual procedure resulted in a set of 124 MedDRA PTs (“the MedDRA psychosis PTs”; listed in ***Supplementary Table 2***) that were used in all analyses requiring psychotic illness to be defined in MedDRA.

### Defining antipsychotics

For most of the analyses of antipsychotics presented in this report that, antipsychotics were defined as the 64 medications in the ATC Level 3 class “Antipsychotics” (listed in ***Supplementary Table 1***). For one analysis – the analysis that defined propsychotics (see below) – a broader set of antipsychotics was defined. The broader antipsychotic set was comprised of (1) the 64 antipsychotics defined using ATC and (2) medications with an indication in the Side Effect Resource (SIDER; version 4.1) – a database which links medications to indications and side effects reported on Food and Medication Administration (FDA) labels^29^ – that mapped to any of the 124 MedDRA psychosis PTs.

### Defining propsychotics

Propsychotics (i.e., medications that induce psychotic symptoms as a side effect) were defined using VigiBase – a medication side effect reporting database with over 15 million reports maintained by the WHO – via the following four-step procedure:

1. VigiBase reports that contained greater than one medication (i.e., reports of side effects that occurred in individuals taking greater than one medication) were removed from the database.
2. Disproportionality analysis^39^ was performed for every medication-side effect link reported in VigiBase as follows. First, a two-by-two table was constructed where rows indicated the number of reports in VigiBase with the medication and columns indicated the number of reports in VigiBase with the side effect. Second, from the values in the two-by-two table, three disproportionality statistics were calculated, each representing a different way of quantifying the amount of evidence in VigiBase suggesting that the medication truly causes the side effect: (a) the number of reports in VigiBase reporting the medication-side effect link, (b) the proportional reporting ratio (PRR), and (c) the Yates’ chi-square test statistic^40,41^.
3. For each medication-side effect link observed in VigiBase, the link was considered true if the following criteria established by other studies^20,21^ were met: (a) at least 3 reports of the medication-side effect link were in VigiBase, (b) the PRR value was greater than or equal to 3, and (c) the Yates’ chi-square test statistic was greater than or equal to 4.
4. Of the medication-side effect links considered true by the previous step, medication-side effects links were filtered out if (a) the medication in the link was in the broad antipsychotic set defined as explained above or (b) the side effect in the link was not one of the 124 MedDRA psychosis PTs. Propsychotics were defined as the 276 medications (linked to 66 psychosis side effects) that remained after applying these filters (***Supplementary Tables 3-4***).

For the analyses that used SIDER to validate the propsychotics identified in VigiBase, propsychotics were defined in SIDER as any medication (after excluding the ATC set of 64 antipsychotics defined above) linked by SIDER to at least one of the 124 MedDRA psychosis PTs. A hypergeometric test was run using R to assess whether the overlap between the two sets of propsychotics was significantly greater than would be expected by chance.

### Defining antipsychotic and propsychotic target genes

#### Data description

Two databases were used to link medications to target genes: DrugBank (version 5.1.1)^23^ and SeaChange (January 13th, 2014 version)^24^. Medication target gene identifiers used in DrugBank and SeaChange were standardized to gene symbols using the UniProt web-based mapping tool^42^. Upon combining DrugBank and SeaChange (i.e., prior to limiting only to medications in VigiBase), 47,985 medication-target gene links were observed (including 2,292 unique medications and 3,417 unique target genes). DrugBank contributed to 10,504 of these medication-target genes links (including 2,156 unique medications and 2,777 unique target genes) and SeaChange contributed to 38,761 of these medication-target gene links (including 792 unique medications and 1,662 unique target genes).

#### Propsychotics

For propsychotics, an empirical p-value was calculated to assess if each prospective target gene (i.e., each target gene linked to >=1 propsychotic) was linked to the propsychotics more than expected by chance. The empirical null distribution used to calculate each of these p-values was generated through 100,000 iterations of the following procedure:

1. A set of medications to randomly sample from was defined (the “background medication set”). The background medication set was the set of medications that (1) were in VigiBase after removing VigiBase reports that included greater than one medication, (2) were not antipsychotics, and (3) could be linked to at least one target gene.
2. A set of 240 medications (i.e., the number of propsychotics linked to >=1 target gene) was randomly selected from the background medication set.
3. The proportion of the randomly selected medications linked to the target gene was recorded.

The empirical p-value was calculated as the fraction of the 100,000 values in the null distribution that were greater than the proportion of propsychotics linked to the target gene. Any target gene with an empirical p-value less than 0.05 was considered a propsychotic target gene.

This procedure for identifying significant propsychotic targets was performed separately for three versions of the data: (1) the full set of 276 propsychotics, with targets from DrugBank and SeaChange; (2) the subset of the 276 propsychotics that remained after removing medications labeled as nervous system drugs by the ATC Level 1 term “Nervous System,” with targets from DrugBank and SeaChange; (3) the full set of 276 propsychotics, with targets from DrugBank only.

#### Antipsychotics

For antipsychotics, an empirical p-value was calculated to assess if each prospective target gene (i.e., each target gene linked to >=1 antipsychotic) was linked to the antipsychotics more than expected by chance. The empirical null distribution used to calculate each of these p-values was generated through 100,000 iterations of the following procedure:

1. A set of medications to randomly sample from was defined (the “background medication set”). The background medication set was the set of medications that (1) were in ATC Level 5, (2) were not propsychotics, and (3) could be linked to at least one target gene.
2. A set of 46 medications (i.e., the number of antipsychotics linked to >=1 target gene) was randomly selected from the background medication set.
3. The proportion of the randomly selected medications linked to the target gene was recorded.

The empirical p-value was calculated as the fraction of the 100,000 values in the null distribution that were greater than the proportion of antipsychotics linked to the target gene. Any target gene with an empirical p-value less than 0.05 was considered an antipsychotic target gene.

### Defining antipsychotic and propsychotic target gene mechanisms of action

#### Data description

DrugBank was used to assign mechanism of action labels to medication-target gene links. Action labels were available for 1,026 propsychotic-target gene links and for 388 antipsychotic-target gene links. Action labels were collapsed into two broad categories: activating (agonist, activator, inducer, potentiator, positive allosteric modulator, positive modulator, and stimulator) and inhibiting (inhibitor, antagonist, blocker, negative modulator, inactivator, suppressor, weak inhibitor, and inhibitory allosteric modulator). For each target gene, the proportion of medications in the class (i.e., propsychotics and antipsychotics) activating and inhibiting the target gene was calculated as the number of medications in the class annotated to the action label divided by the total number of medications in the class with any mechanism data in DrugBank (***Supplementary Table 8***). The significance of these proportions was assessed using the following permutation procedures.

#### Propsychotics

Two empirical values were calculated for each propsychotic target gene with a mechanism of action label: one empirical p-value to assess whether the target gene was assigned an activating mechanism of action label more than expected by chance and one empirical p-value to assess whether the target gene was assigned an inhibiting mechanism of action label more than expected by chance. The empirical null distributions used to calculate these p-values was generated through 100,000 iterations of the following procedure:

1. A set of medications to randomly sample from was defined (the “background medication set”). The background medication set was the set of medications that (1) were in VigiBase after removing VigiBase reports that included greater than one medication, (2) were not antipsychotics, and (3) could be linked to at least one target gene with a mechanism of action label.
2. A set of 167 medications (i.e., the number of propsychotics linked to >=1 target gene with a mechanism of action label) was randomly selected from the background medication set.
3. The proportion of the randomly selected medications linked to the target gene with an activating mechanism of action label was recorded.
4. The proportion of the randomly selected medications linked to the target gene with an inhibiting mechanism of action label was recorded.

The empirical p-value for the activating mechanism of action label was calculated as the fraction of the 100,000 values in the corresponding null distribution (i.e., the null distribution created from the values in the third step of the permutation procedure) that were greater than the proportion of propsychotics linked to the target gene with an activating mechanism of action. Any target gene with an empirical p-value less than 0.05 was considered a propsychotic target gene with an activating mechanism of action.

The empirical p-value for the inhibiting mechanism of action label was calculated as the fraction of the 100,000 values in the corresponding null distribution (i.e., the null distribution created from the values in the fourth step of the permutation procedure) that were greater than the proportion of propsychotics linked to the target gene with an inhibiting mechanism of action. Any target gene with an empirical p-value less than 0.05 was considered a propsychotic target gene with an inhibiting mechanism of action.

#### Antipsychotics

Two empirical values were calculated for each antipsychotic target gene with a mechanism of action label: one empirical p-value to assess whether the target gene was assigned an activating mechanism of action label more than expected by chance and one empirical p-value to assess whether the target gene was assigned an inhibiting mechanism of action label more than expected by chance. The empirical null distributions used to calculate these p-values was generated through 100,000 iterations of the following procedure:

1. A set of medications to randomly sample from was defined (the “background medication set”). The background medication set was the set of medications that (1) were in ATC Level 5, (2) were not propsychotics, and (3) could be linked to at least one target gene with a mechanism of action label.
2. A set of 44 medications (i.e., the number of antipsychotics linked to >=1 target gene with a mechanism of action label) was randomly selected from the background medication set.
3. The proportion of the randomly selected medications linked to the target gene with an activating mechanism of action label was recorded.
4. The proportion of the randomly selected medications linked to the target gene with an inhibiting mechanism of action label was recorded.

The empirical p-value for the activating mechanism of action label was calculated as the fraction of the 100,000 values in the corresponding null distribution (i.e., the null distribution created from the values in the third step of the permutation procedure) that were greater than the proportion of antipsychotics linked to the target gene with an activating mechanism of action. Any target gene with an empirical p-value less than 0.05 was considered an antipsychotic target gene with an activating mechanism of action.

The empirical p-value for the inhibiting mechanism of action label was calculated as the fraction of the 100,000 values in the corresponding null distribution (i.e., the null distribution created from the values in the fourth step of the permutation procedure) that were greater than the proportion of antipsychotics linked to the target gene with an inhibiting mechanism of action. Any target gene with an empirical p-value less than 0.05 was considered an antipsychotic target gene with an inhibiting mechanism of action.

### Enrichment of antipsychotic and propsychotic target genes for psychosis risk genes

The strength of association between psychosis risk and a gene was defined in two ways: (1) “rare LoF variant psychosis risk” and (2) “common SNP psychosis risk.” To define rare LoF variant psychosis risk, a summary statistics file from the SCHEMA study was downloaded (i.e., Supplementary Table 5 from the SCHEMA study publication)^17^. This file was processed to (1) remove genes with no value in the “P meta” column, (2) remove genes appearing in greater than one row of the file, and (3) retain genes that were the target gene of at least one medication in DrugBank or SeaChange. The remaining 2,692 SCHEMA p-values defined rare LoF variant psychosis risk. To define common SNP psychosis risk, summary statistics from the PGC3SCZ GWAS were downloaded from the URL provided in the PGC3SCZ GWAS publication (the version of the summary statistics used was the file called “PGC3_SCZ_wave3.primary.autosome.public.v3.vcf.tsv.gz”)^15^. SNPs appearing in more than one row of the summary statistics file were removed, and the resulting summary statistics file was uploaded to the FUMA web server (https://fuma.ctglab.nl; accessed April 4^th^, 2024)^43^. Using the SNP2GENE tool on the FUMA web server, gene-level p-values were calculated from the SNP summary statistics. The default SNP2GENE tool parameter settings were used with two exceptions: (1) the “Perform MAGMA” option was selected under the “MAGMA Analysis” section of the job submission form; (2) the “Perform eQTL mapping” option was selected under the “Gene Mapping (eQTL mapping)” section of the job submission form, and “GTEx v8 Brain (13)” was selected as the “Tissue Type” value to be used in the eQTL mapping analysis. Default SNP2GENE tool parameter settings included the reference panel population set to “1000G Phase3 EUR” and the exclusion of the major histocompatibility region. The PGC3SCZ GWAS gene-level p-values used in downstream analyses were contained in the “magma.genes.out” file in the output of the job submission (column titled “P”). This file was processed to (1) remove genes with more than one result and (2) retain genes that were the target gene of at least one medication in DrugBank or SeaChange. The remaining 2,621 p-values defined common SNP psychosis risk. The two types of psychosis genetic risk (i.e., rare LoF variant psychosis risk and common SNP psychosis risk) were tested for enrichment in two target gene sets (i.e., propsychotic target genes and antipsychotic target genes) using the following procedure: (1) the negative log_10_ value was calculated for each psychosis genetic risk p-value; (2) the distribution of the negative log_10_ p-values of target genes was compared to the distribution of the negative log_10_ p-values of all other genes using the wilcox.test() function of the base stats R package, with the alternative hypothesis specifying the expectation that target genes would have more significant p-values than other genes. High-confidence psychosis risk genes were defined in SCHEMA as the 10 genes with “P meta” values below the exome-wide significance threshold defined by the SCHEMA authors (2.14 x 10^-6^). High-confidence psychosis risk genes were defined in the PGC3SCZ GWAS as genes with Bonferroni-adjusted MAGMA p-values below 0.05. Adjustment of the MAGMA p-values was performed in R using the p.adjust() function of the base stats R package.

### GRIN2A case report

Genetic and clinical data from the Mount Sinai Million Health Discoveries Program (MSM-HDP; formerly, the BioMe Biobank Program; N = 29,064) were analyzed to identify carriers of rare LoF variants in *GRIN2A* with psychotic illness. MSM-HDP study activities for the current report were approved by the Icahn School of Medicine at Mount Sinai’s Institutional Review Board (Institutional Review Board 07–0529) and all study participants provided written informed consent. Data analyzed from MSM-HDP participants has been previously described, including pipelines for variant calling and quality control^44,45^. The genetic data was comprised of DNA sequence variants identified through whole-exome sequencing. DNA sequence variants identified in MSM-HDP participants were annotated using a workflow modeled after the workflow used in SCHEMA^46^. Specifically, annotation by LOFTEE (as implemented in the Variant Effect Predictor)^47^ was applied to variants that passed quality control filters. LoFs were defined as any variant annotated by the LOFTEE plugin as “loss-of-function” with either “high confidence” or “low confidence.” LoFs were defined as rare if the minor allele count in the MSM-HDP cohort was less than or equal to five. The electronic medical records of carriers in MSM-HDP of rare LoFs in *GRIN2A* defined in this manner were reviewed by a study psychiatrist (AWC). The individual with evidence of psychotic illness (i.e., the founder) was recalled by the study team to obtain further details on the history of psychotic illness.

## Supplementary Information

### Standardizing clinical concept terminology in VigiBase

#### Overview

The reports in the version of the VigiBase database analyzed for this study contained (1) a description of the side effect that arose in the subject of the report as a result of a medication the subject was taking and (2) descriptions of conditions for which the subject of the report was being treated (i.e., indications) at the time of the side effect. The side effect descriptions were provided already standardized to MedDRA codes, but indication descriptions were not provided in a single standardized terminology. For example, the indication schizophrenia had over 50 unique representations in the database (e.g., “Schizophrenia NOS”, “Other schizophrenia”, “Schizophrenia, unspecified”, “Schizophrenia, undifferentiated type”, “Unspecified schizophrenia, unspecified state”). To perform analyses on VigiBase data that consider information about indications, it was necessary to indication terms to MedDRA. This was accomplished through a multistep procedure described in this section that incorporated a variety of medical lexicon mapping databases through both exact and partial string matching. To facilitate mapping across lexicons, strings were pre-processed prior to mapping using the following four-step procedure: (1) letters were made lowercase; (2) whitespaces and special characters were converted into a period; (3) instances of 2 or more consecutive periods were converted to a single period; (4) leading and trailing whitespaces were removed.

#### Exact string matching

Exact string matching was used to map VigiBase indications to MedDRA terms in the MedDRA source files (version 20.0). Unmapped indications were then mapped using exact string matching to the Unified Medical Language System (UMLS, version 2016AB). The UMLS version used included MedDRA terms (version 19.0), and UMLS Concept Unique Identifiers (CUI) map MedDRA terms to other lexicons. Remaining unmapped VigiBase indications were then mapped using exact string matching to the Observational Medical Outcomes Partnership (OMOP) Common Data Model, which like the UMLS connects MedDRA terms to terms in other medical lexicons.

#### Partial string matching

For all VigiBase indications that could not be mapped to MedDRA using exact string matching, a partial string-matching approach was utilized. Two partial string-matching tools were utilized in this procedure: Usagi and Fuzzywuzzy. The Observational Health Data Sciences and Informatics (OHDSI) Usagi software (v1.0.0x)^48^ is a partial string mapping tool for bioinformatics that has been used by other studies to map medication names to RxNorm^49^. Usagi performs matching utilizing the Apache Lucene library, which is a suite of tools commonly used for computer programming tasks that require text searching. In the case of Usagi, the texts that are searched are the suite of medical terminology databases (e.g., RxNorm, MedDRA) that are contained in the OHDSI data files, and the terms searched for are those input by the Usagi user (e.g., VigiBase indications). The Usagi software pre-processes the user-supplied terms with an implementation of the Porter stemmer algorithm that reduces words to their stems (e.g., “adults” becomes “adult”). For the current task, Usagi was run with MedDRA as the text to search and using as input unmapped VigiBase indications. For each term, Usagi outputs the single best match along with a matching confidence score.

In addition to Usagi, the python library fuzzywuzzy was used for partial string mapping. Fuzzywuzzy contains a suite of algorithms for term similarity matching that are all based on edit distance, which is the minimum number of operations that are required to transform one string into the other. Given two strings, *i* and *j*, fuzzywuzzy calculates the edit distance as 2*(number of elements shared by *i* and *j*)/(number of elements in *i* plus the number of elements in *j*). What differentiates the algorithms in fuzzywuzzy from one another is how the two strings being compared are processed prior to performing this calculation. For the current task, four of the algorithms in the fuzzywuzzy package were utilized: WRatio, QRatio, token_set_ratio, and token_sort_ratio. The QRatio algorithm calculates edit distance on unprocessed strings. This approach fails when the two strings compared consist of the same words in different orders, for instance when comparing “hello world” to “world hello.” In the case of VigiBase indications, this algorithm would not, for example, recognize these two indications as the same concept: “Schizophrenia, undifferentiated” and “Undifferentiated schizophrenia.” The token_sort_ratio algorithm performs well with such instances, as it splits the strings into composite “tokens” (in this case, words) and compares the two strings to one another after sorting the tokens (e.g., alphabetically). The token_set_ratio algorithm builds upon the token_sort_ratio algorithm by further considering differences between the two strings being compared that might artificially make the two strings appear different when in fact they contain the same information. For example, consider these two potential strings that could appear in the VigiBase indications: “the condition the patient suffered from was schizophrenia” and “schizophrenia.” Using the token_sort_ratio algorithm, the similarity score for these two terms would be low since many letters would have to be added to “schizophrenia” to make it equal the first string. The token_set_ratio algorithm calculates the edit distance between the tokens that overlap with one another in the strings (in this case, “schizophrenia”), then generates three values by adding to this value the number of characters in the remaining tokens of string 1, string 2, and the combination of string 1 and string 2. It then outputs the highest of these three values as the similarity score. Since in this example string 2 has no additional characters, the maximum score would be based on the edit distance between the “schizophrenia” and “schizophrenia” (which is 0), and therefore a high score would result, which would be ideal since in fact these two values are, for the purposes of this task, the same clinical concept. The WRatio algorithm is not a unique algorithm in itself, but rather is a process by which all of the fuzzywuzzy algorithms are run and the outputs are weighted, and the highest weighted score returned. The four fuzzywuzzy algorithms were run on all unmapped VigiBase indications. In contrast to Usagi, the data files that fuzzywuzzy uses to match terms against are supplied by the user. Therefore, for this mapping procedure the input against which unmapped VigiBase indications were matched was a list of MedDRA codes from the MedDRA source files.

Having run five different partial string-matching algorithms for each of the VigiBase indications that could not be mapped using exact string matching, it was then necessary to assess the performance of the algorithms. For every string match identified by one of the algorithms, the algorithm outputs a confidence score for the match. While these scores are informative in many respects, the performance of these algorithms depends on the nature of the input terms. Therefore, a manual inspection procedure was carried out to establish confidence score cutoffs for each algorithm below which string matches would be discarded. Matches were randomly sampled in batches of 10 from each algorithm’s output at confidence score windows of size 0.1 (starting at 0.5) and up to 50 terms were evaluated per algorithm manually by a study physician (AWC) for correctness. For all five partial string-matching algorithms used, the performance of the algorithm in a given window was quantified as the proportion of the matches in the window that were manually reviewed that were scored by the reviewer as a true match. For each algorithm, a threshold was selected that maximized the number of true matches identified. In all, over 90% of the initial VigiBase indication terms were mapped to a MedDRA code through the exact and partial string-matching procedures.

### Standardizing medication names in VigiBase

VigiBase medication identifiers were provided in terms defined in the WHO Medication Dictionary (WDD; version of December 1st, 2016). In the WDD, medicinal products are mapped to a “Substance ID” that represents the substances in the product. Two WDD medicinal products with the same active substance (e.g., two salt forms of the same medication) therefore have different Substance IDs. For instance, the medication naproxen appears in the WDD both as “naproxen” (WDD Substance ID 6340) and “naproxen sodium” (WDD Substance ID 6653). Connecting these two Substance IDs to one another is not possible using the WDD but is crucial to link VigiBase medication names to medication names used in other databases analyzed in this report and (2) calculate accurate disproportionality analyses statistics for this report. When the analyses for this report were performed, official maps between WDD and RxNorm codes had not been created by either the WDD or RxNorm developers, so it was necessary to create such a map for this report.

The mapping procedure began with two text identifiers for each medication in VigiBase: the substance name (e.g., “naproxen”) and, for single-ingredient medications, the product name (e.g., “naprometin”). To facilitate mapping across lexicons, strings were pre-processed prior to mapping using the following four-step procedure: (1) letters were made lowercase; (2) whitespaces and special characters were converted into a period; (3) instances of 2 or more consecutive periods were converted to a single period; (4) leading and trailing whitespaces were removed. Both exact and partial string-matching methods were then utilized to map WDD to RxNorm. Exact string matching was performed first. A manually curated list of words deemed uninformative for this study’s purposes (e.g., salts and descriptors) were removed from WDD strings prior to exact string matching. Next, a partial string-matching algorithm provided by RxNorm developers as an API was used to map the remaining unmapped WDD terms to RxNorm RXCUI^50^. Each match output by this algorithm is given a confidence score from 1 to 100. Manual evaluation of these scores and the quality of the matches found that scores above 50 returned correct matches nearly 100% of the time, and all matches with a score above 50 were called true matches. For matches with scores less than 50, true matches were called if the following criteria were met (these criteria were determined by manual investigation): (1) the first word of the WDD and RxNorm terms in the match were identical to one another; (2) the first word of the WDD and RxNorm terms in the match were not salt names; (3) >50% of the words in the WDD term represented >50% of the words in the RxNorm term. After applying these exact and partial string-matching procedures, approximately 80% of WDD names could be mapped to at least one RxNorm RXCUI.

### Standardizing medication names in SIDER

SIDER medications are provided in the form of STITCH codes^51^. When the analyses for this report were performed, there was no official map between STITCH codes and RxNorm codes provided by either the STITCH source files or the RxNorm source files. Therefore, a map between these lexicons had to be created for this report. Six mapping pathways were devised, and each is detailed in this section. Altogether, employing these six mapping pathways results in over 95% of the STITCH codes in SIDER mapping to at least one RxNorm RXCUI.

#### STITCH to RxNorm Mapping Pathway 1

While the STITCH source files did not contain mappings between STITCH codes and RxNorm codes, they did contain mappings between STITCH codes and the medication codes used by other lexicons. Some of these other lexicons were also in the RxNorm source files (e.g., ATC, DrugBank). By utilizing the lexicons present in both the STITCH source files and the RxNorm source files as intermediates, many STITCH codes in SIDER could be mapped to RxNorm RXCUI.

#### STITCH to RxNorm Mapping Pathway 2

STITCH codes were mapped to compound names in the STITCH source files. Compound names were mapped to the string name in RxNorm source files using exact string matching. RxNorm strings were mapped to RxNorm RXCUI using the RxNorm source files.

#### STITCH to RxNorm Mapping Pathway 3

To map the remaining unmapped STITCH codes to RxNorm RXCUIs required leveraging the fact that some of the lexicons linked to STITCH codes in the STITCH source files that were not present in the RxNorm source files could be mapped to lexicons in RxNorm using a mapping procedure that incorporated information from additional databases. Four such mapping procedures were identified (i.e., STITCH to RxNorm Mapping Pathways 3-6), and the first of these four was as follows. STITCH codes were mapped to PubChem compound identifiers (CIDs) in the STITCH source files. PubChem CIDs were linked to Food and Medication Administration (FDA) Unique Ingredient Identifier (UNII) codes UNII through (a) a linker file provided by PubChem (Linker File 1; the URL used to download this file is provided below) and (b) a linker file provided by UniChem (Linker File 2; the URL used to download this file is provided below). UNII codes could be linked to RxNorm RXCUI by a linker file provided by the FDA Substance Registration System (SRS) (Linker File 3; the URL used to download this file is provided below).

#### STITCH to RxNorm Mapping Pathway 4

STITCH codes were mapped to PubChem CIDs in the STITCH source files. PubChem CIDs were mapped to FDA Structured Product Labeling (SPL) codes using the PCIES website. SPL codes were mapped to RxNorm RXCUI using the UMLS (version 2016AB).

#### STITCH to RxNorm Mapping Pathway 5

STITCH codes were mapped to FDA National Drug File (NDF) codes using a PubChem source file (Linker File 4). NDF codes were mapped to RxNorm RXCUIs using UMLS.

#### STITCH to RxNorm Mapping Pathway 6

STITCH codes were mapped to PubChem CIDs in the STITCH source files. PubChem CIDs were mapped to Simplified Molecular Input Line Entry System (SMILES) codes using the PCIES website. SMILES were mapped to RxNorm RXCUIs using Linker File 3.

Linker File 1 URL: https://pubchem.ncbi.nlm.nih.gov/source/2322#data=Annotations

Linker File 2 URL: ftp://ftp.ebi.ac.uk/pub/databases/chembl/UniChem/data/wholeSourceMapping/src_id14/src14src22.txt.gz

Linker File 3 URL: https://fdasis.nlm.nih.gov/srs/download/srs/UNII_Data.zip

Linker File 4 URL: https://ftp.ncbi.nlm.nih.gov/pubchem/RDF/compound/general/pc_compound_type.ttl.gz

PCIES website:

https://pubchem.ncbi.nlm.nih.gov/docs/identifier-exchange-service

### Standardizing DrugBank Medication Names

The medication codes provided in DrugBank (version 5.1.1) are codes created by the DrugBank developers specifically for DrugBank. Since DrugBank is one of the lexicons in the RxNorm source files, DrugBank codes could be mapped to RxNorm RXCUI through the RxNorm source files. Of note, only approximately one third of the DrugBank codes mapped to a RxNorm RXCUI with a SAB value of “RXNORM”. This is because many of the medication compounds in DrugBank are not medications approved for use in humans. Filtering DrugBank codes for those that are medications approved for clinical use (based data provided in the DrugBank source files), the RxNorm source files successfully mapped over 90% of DrugBank medications approved for clinical use to an RXCUI with SAB value of “RXNORM”.

### Standardizing medication names in SeaChange

In the SeaChange source files provided to the research team by the SeaChange developers, medications were identified as ChEMBL codes^52^. Since ChEMBL was not one of the lexicons in the RxNorm source files used for this report, a map between ChEMBL codes and RxNorm RXCUIs was created using the following procedure.

1. Using ChEMBL source files (accessed via the URLs below), the “preferred name” values for the ChEMBL codes in SeaChange were identified, and exact string matching was used to map these preferred names to medication names in RxNorm
2. Using ChEMBL source files and UniChem source files (accessed via the URLs below), ChEMBL codes in SeaChange were linked to codes of other medication lexicons (e.g., UNII, PubChem) that could then be linked to RxNorm RXCUIs using the RxNorm source files
3. Using Linker File 3 (see above), ChEMBL codes in SeaChange were mapped to RxNorm RXCUIs

Using these three strategies, approximately 75% of the ChEMBL codes in SeaChange could be mapped to at least one RxNorm RXCUI. Manual inspection of the unmappable codes revealed that most represented compounds with no approved clinical indications in humans.

#### ChEMBL source files and UniChem source files

ftp://ftp.ebi.ac.uk/pub/databases/chembl/UniChem/data/wholeSourceMapping/src_id1/src1src14.txt.gz

ftp://ftp.ebi.ac.uk/pub/databases/chembl/UniChem/data/wholeSourceMapping/src_id1/src1src7.txt.gz

ftp://ftp.ebi.ac.uk/pub/databases/chembl/UniChem/data/wholeSourceMapping/src_id1/src1src2.txt.gz

ftp://ftp.ebi.ac.uk/pub/databases/chembl/UniChem/data/wholeSourceMapping/src_id1/src1src22.txt.gz

ftp://ftp.ebi.ac.uk/pub/databases/chembl/UniChem/data/wholeSourceMapping/src_id1/src1src4.txt.gz

ftp://ftp.ebi.ac.uk/pub/databases/chembl/UniChem/data/wholeSourceMapping/src_id2/src2src22.txt.gz

ftp://ftp.ebi.ac.uk/pub/databases/chembl/ChEMBLdb/latest/chembl_22_1_chemreps.txt.gz

ftp://ftp.ebi.ac.uk/pub/databases/chembl/ChEMBLdb/latest/chembl_uniprot_mapping.txt

### Standardizing medication names in ATC

Medications in the ATC source files were provided as ATC codes. Level 5 ATC codes in the ATC source files were mapped to RxNorm RXCUIs using the RxNorm source files since ATC was one of the lexicons in the RxNorm version used for this report.

### Standardizing RxNorm RXCUIs

For the purposes of the current study, different RxNorm RXCUIs could represent the same medication. Therefore, after procedures described above for standardizing medication names to RxNorm were completed, it then was necessary to relate RxNorm RXCUIs to one another by finding the RxNorm RXCUI representative of active ingredients. The RxNorm API was used to (1) map RxNorm RXCUIs to “IN” values and (2) obtain the “status” of each RxNorm RXCUI in the RxNorm release active at the time the RxNorm API was accessed. Possible status values for a RxNorm RXCUI in the RxNorm API are “Active” (the concept is in the current RxNorm data set and has a non-obsolete term from the RxNorm lexicon), “Alien” (the concept exists in the current RxNorm data set, but contains only terms from lexicons other than RxNorm), “Quantified” (the concept is inactive and has been quantified to one or more concepts in the current RxNorm data set), “Remapped” (the concept has been remapped to one or more concepts in the current RxNorm data set), “Retired” (the concept no longer exists in the current RxNorm data set, or contains only obsolete terms), or “Unknown” (the concept identifier is invalid). Only RxNorm RXCUIs with Active status were retained.

## Data Availability

Publicly available data analyzed for this report was obtained through the following URLs:
VigiBase: https://who-umc.org/vigibase/.
SIDER: http://sideeffects.embl.de/
DrugBank: https://go.drugbank.com/
RxNorm: https://www.nlm.nih.gov/research/umls/rxnorm/index.html
MedDRA: https://www.meddra.org/
SCHEMA: https://schema.broadinstitute.org/
PGC3SCZ: https://pgc.unc.edu/for-researchers/download-results/

## Supplementary table captions

**Supplementary Table 1**

List of the 64 antipsychotic drugs defined using the Anatomical Therapeutic Chemical Classification (ATC) Level 3.

**Supplementary Table 2**

List of the 124 Medical Dictionary for Regulatory Activities (MedDRA) terms used to represent psychotic illness.

**Supplementary Table 3**

List of the 276 propsychotic drugs defined using Vigibase.

**Supplementary Table 4**

List of 66 adverse events associated with the propsychotic drugs in Supplementary Table 3.

**Supplementary Table 5**

The proportion of propsychotics mapping to each of 85 different pharmacological subgroups defined by ATC Level 3.

**Supplementary Table 6**

Results of analyses linking propsychotics and antipsychotics to target genes.

**Supplementary Table 7**

Results of analyses linking propsychotics and antipsychotics to mechanisms of action on target genes.

**Supplementary Table 8**

Table of targets showing the observed proportion and p-value for upregulated and downregulated mechanisms of action.

## Data and materials availability

Publicly available data analyzed for this report was obtained through the following URLs:

- VigiBase: https://who-umc.org/vigibase/.
- SIDER: http://sideeffects.embl.de/
- DrugBank: https://go.drugbank.com/
- RxNorm: https://www.nlm.nih.gov/research/umls/rxnorm/index.html
- MedDRA: https://www.meddra.org/
- SCHEMA: https://schema.broadinstitute.org/
- PGC3SCZ: https://pgc.unc.edu/for-researchers/download-results/

All code used for this report will be made available upon publication.

